# Longitudinal Filovirus Seroreactivity Following Ad26.ZEBOV (Zabdeno) and MVA-BN-Filo (Mvabea) Vaccination in the Democratic Republic of the Congo

**DOI:** 10.64898/2026.10.15.26365698

**Authors:** Hannah L. Wallace, Megan Halbrook, Jean Paul Kompany, Nicole A. Hoff, Sydney Merritt, Patrick K. Mukadi, Michael Beya, Handdy Kalengi, Merly Tambu, Zéphi Mossoko, J. Daniel Kelly, Aquena H. Ball, Albert To, Teri Wong, Hugo Kavunga, Lisa E. Hensley, Axel T. Lehrer, Jason Kindrachuk, Placide Mbala-Kingebeni, Anne W. Rimoin

**Author notes:** Corresponding author: Anne W. Rimoin. Contributed equally as first authors. Joint Senior Authors (Jason Kindrachuk, Placide Mbala-Kingebeni, Anne W. Rimoin).

## Abstract

**Background:** Filoviruses, specifically Ebola virus (EBOV), Bundibugyo virus (BDBV), and Marburg virus (MARV), have been the etiologic agents of multiple outbreaks of severe disease in the Democratic Republic of the Congo (DRC). Two EBOV-specific vaccine regimens have been approved for use in outbreak scenarios, including the Ad26.ZEBOV/MVA-BN-Filo (Zabdeno/Mvabea) two-dose vaccine, where the MVA-BN-Filo booster dose contains EBOV, MARV, and Sudan (SUDV) virus glycoproteins (GPs) and the nucleoprotein from Tai Forest virus. While long-term immunogenicity of up to two years has been found in clinical trial settings, limited real-world data exists.

**Methods:** Using a multiplexed immunoassay, samples (*n*=698 at baseline) collected in Mbandaka, Equateur province, DRC, between September 2023 and October 2024 were assessed for seroreactivity to six filovirus antigens (1 BDBV, 2 EBOV, 2 MARV, and 1 SUDV) at three distinct periods: baseline/pre-vaccination, 6-8 weeks post-first dose (Zabdeno), at booster administration (Mvabea), and one-year post-boost dose.

**Results:** Substantial filovirus seroreactivity was found at baseline, with 48.7% (*n*=340/698) of individuals being seroreactive to at least one virus antigen. High baseline seroreactivity was found against BDBV GP (31.1%, *n*=217/698), with lower baseline seroreactivity to EBOV GP (15.5%, *n*=106/698), comparatively. Overall EBOV GP seroreactivity increased to 66.8% (*n*=376/563) at one year-post vaccine boost and 57.6% (*n*=250/434) of participants were seroreactive. Despite the booster dose inclusion of SUDV GP and MARV GP, there were no substantial increases in reactivity against either antigen at subsequent timepoints. Overall filovirus seroreactivity increased at all timepoints for the entire cohort, although filovirus reactivity decreased over the study period for those who were seroreactive at baseline.

**Findings:** These data highlight detection of significantly elevated seoreactivity to EBOV GP following first-dose administration of Ad26.ZEBOV, which remained relatively stable one-year post-booster dose administration. Additionally, this study identifies high levels of baseline seroreactivity to BDBV GP despite no previous documented outbreaks in this region of DRC. These data suggest that vaccination with the Ad26.ZEBOV-MVA-BN-Filo vaccine induces stable seroreactivity against EBOV GP, but not to other filovirus antigens.

## Introduction

The recent reemergence of Bundibugyo virus (BDBV) in April 2026 in Ituri Province, Democratic Republic of the Congo (DRC) prompted an almost immediate declaration of a Public Health Emergency of International Concern by the World Health Organization. ^1^ Given the rapid geographic expansion of the outbreak to multiple regions in Eastern DRC and Uganda, with 9,212 confirmed infections and 4,441 fatalities as of 15 October 2026 ^2^, strengthening understanding of potential medical countermeasures to this high-consequence pathogen is paramount. There are currently no licensed or approved vaccines for use during BDBV outbreaks.

BDBV is a member of the *Filoviridae* family, a group of single-stranded, negative-sense RNA viruses associated with significant morbidity and mortality. Beyond BDBV, Ebola virus (EBOV), Sudan virus (SUDV), and Marburg virus (MARV) have also been associated with multiple Filovirus disease (FVD) outbreaks. The DRC has recorded eighteen unique FVD outbreaks to date, with three in Equateur (EQT) province specifically since 2018.^3^ Due to repeated exposures to filoviruses in specific regions, questions have been raised regarding population-level immunity and potential cross-protection in the context of future FVD outbreaks.

As EBOV has been most frequently associated with outbreaks and high fatality rates, vaccine development has historically focused on this virus species. Two vaccines have been approved for individuals at risk of EBOV infection, the single dose rVSVΔG-ZEBOV-GP vaccine (Ervebo), and the heterologous Ad26.ZEBOV (Zabdeno), MVA-BN-Filo (Mvabea)^1^ vaccine (hereafter referred to as Ad26.ZEBOV-MVA-BN-Filo). The heterologous two-dose vaccine includes a monovalent replication-incompetent adenoviral vector serotype 26 that encodes the full-length EBOV glycoprotein (GP) (Ad26.ZEBOV), and a non-replicating multivalent modified vaccinia Ankara (MVA) encoding the GPs of EBOV, SUDV, and MARV and the Tai Forest virus nucleoprotein (NP; MVA-BN-Filo).^4^ Importantly, the MVA boost does not encode for any BDBV proteins.

Data from rVSVΔG-ZEBOV-GP ring vaccination trials during the 2013-2016 West African EBOV epidemic and during EVD outbreaks in the DRC have suggested that the vaccine is highly efficacious.^5–9^ While Ad26.ZEBOVMVA-BN-Filo vaccination has been shown to be safe, highly immunogenic, and efficacious in non-human primate models^10^, efficacy data is not available from human clinical studies.^11,12^ Therefore, vaccine effectiveness against EBOV or other filoviruses that cause disease in humans is unclear. While data suggests that anti-EBOV GP antibodies cross-react with BDBV, the extent to which this translates into meaningful protection immune responses and protection against BDBV disease remains uncertain.^13–17^ Additionally, long-term studies of vaccine efficacy and the effect of vaccination in populations with pre-existing immunity or prior exposures in endemic settings are lacking. Thus, we evaluated seroreactivity against multiple filovirus antigens from BDBV, EBOV, MARV, and SUDV in a Ad26.ZEBOV-MVA-BN-filo vaccinated cohort in DRC sampled longitudinally with collection prior to vaccination, eight weeks post-first dose/at booster administration, and one-year post-booster vaccination in Equateur (EQT) province.

## Methods

### Participants and procedures

This cohort study recruited adults from Wangata, Bolenge, and Mbandaka health zones in EQT, DRC between September 2023 and October 2024. Samples included in this analysis were collected as part of a larger study, targeting areas of the DRC that have historically experienced filovirus outbreaks. All participants were 18 years or older and at each visit, and screened for fever by trained personnel. Prior to blood draw, participants were asked about any symptoms associated with recent or acute illness. Individuals experiencing illness at the time of collection and those who had reported prior EBOV vaccination (rVSVΔG-ZEBOV-GP) were excluded from the study. Participants were administered a tablet-based survey, underwent a brief physical examination by trained field staff, and provided a blood sample collected in red top serum tubes. Blood samples were centrifuged, and serum was heat inactivated for 30 minutes at 56°C for transport and storage; 1 mL serum aliquots were stored at -20°C in field laboratories and transferred to -80°C for long-term storage in Kinshasa, DRC. Blood samples were collected over three visits.

### Serological Analyses

Antibody testing was performed at the Institut National de Researche Biomédicale (INRB), in Kinshasa, DRC, using a custom pan-filovirus multiplex immunoassay (MIA) developed at the University of Hawaii.^18^ This MIA provides the mean fluorescence intensity (MFI) of reactivity against EBOV GP, EBOV viral protein 40 (VP40), BDBV GP, SUDV GP, MARV GP and MARV VP40. The selection and production of the antigens and assay protocols have been described elsewhere.^14^ Briefly, serum samples were diluted 1:100 and the assay performed in duplicate with background determination using bovine serum albumin (BSA). Prior to analysis, background reactivity to BSA was subtracted from the individual antibody reactivity MFI values. Seroreactivity thresholds specific to the DRC were previously determined.^19^ Reactivity BSA-adjusted MFI thresholds were as follows: EBOV GP, 2589; EBOV VP40, 10751; BDBV GP, 4433; SUDV GP, 9554; MARV GP, 4828; and MARV VP40, 9203. Binary seroreactivity for any antigen at any timepoint was BSA-adjusted MFI above that of the thresholds for each antigen.

### Subgroup Analyses

Various subgroup analyses were undertaken, including stratifying the data by overall reactivity (those with reactivity to at least one antigen) at baseline (visit 0), those who attended all three study visits, sex, age, and health care provider status as all of these factors can either influence immune responses or change individual risk factors related to previous filovirus exposure. Individuals with EBOV GP-specific reactivity at baseline were removed for subgroup analyses to alleviate the effect of previous filovirus exposure and/or previous vaccination to allow for identification of changes to reactivity among a naïve population.

### Data visualization and statistical analyses

Data was analyzed using R Studio v4.5.0 (version 4.5.2, R Core Team, 2026) and Microsoft Excel. Figures generated using Biorender software have been appropriately noted. Proportions of individuals with overall or specific relativity were compared using a two-sample test for equality of proportions with continuity correction and ANOVAs were used to compare MFI variance between groups with Tukey’s post-hoc tests used to identify where significant differences were observed.

### Ethical Approvals

This study was approved by the Institutional Review Board at the University of California, Los Angeles (IRB#20-001091) as well as by the Ethics Committee at the Kinshasa School of Public Health at the University of Kinshasa (ESP/CE/142/2020). All samples were de-identified prior to analysis.

## Results

### Study Population Demographics and Baseline Seroreactivity

For this study, 698 participants were enrolled, of which 673 individuals (96·4%) had available demographic data (visit 0; **Figure 1**). Blood samples were subsequently collected over three visits with the visit 0 preceding the first administration of the Ad26.ZEBOV vaccine. The MVA-BN-Filo booster vaccine was administered at the second visit (visit 1), six to eight weeks post-visit 0. The final visit (visit 2) occurred one-year following MVA-BN-Filo booster administration (visit 1). The cohort included significantly more male (62%, *n*=421/673) than female (38%, *n*=253/673) study participants. Participant age ranged from 18 to 85, with a mean age of 30.8 and 38 individuals identified as health care providers (5·6%). At visit 1, 563 (80·7%) individuals participated and at visit 2, 434 (62·2%) individuals participated.

**Figure 1.**
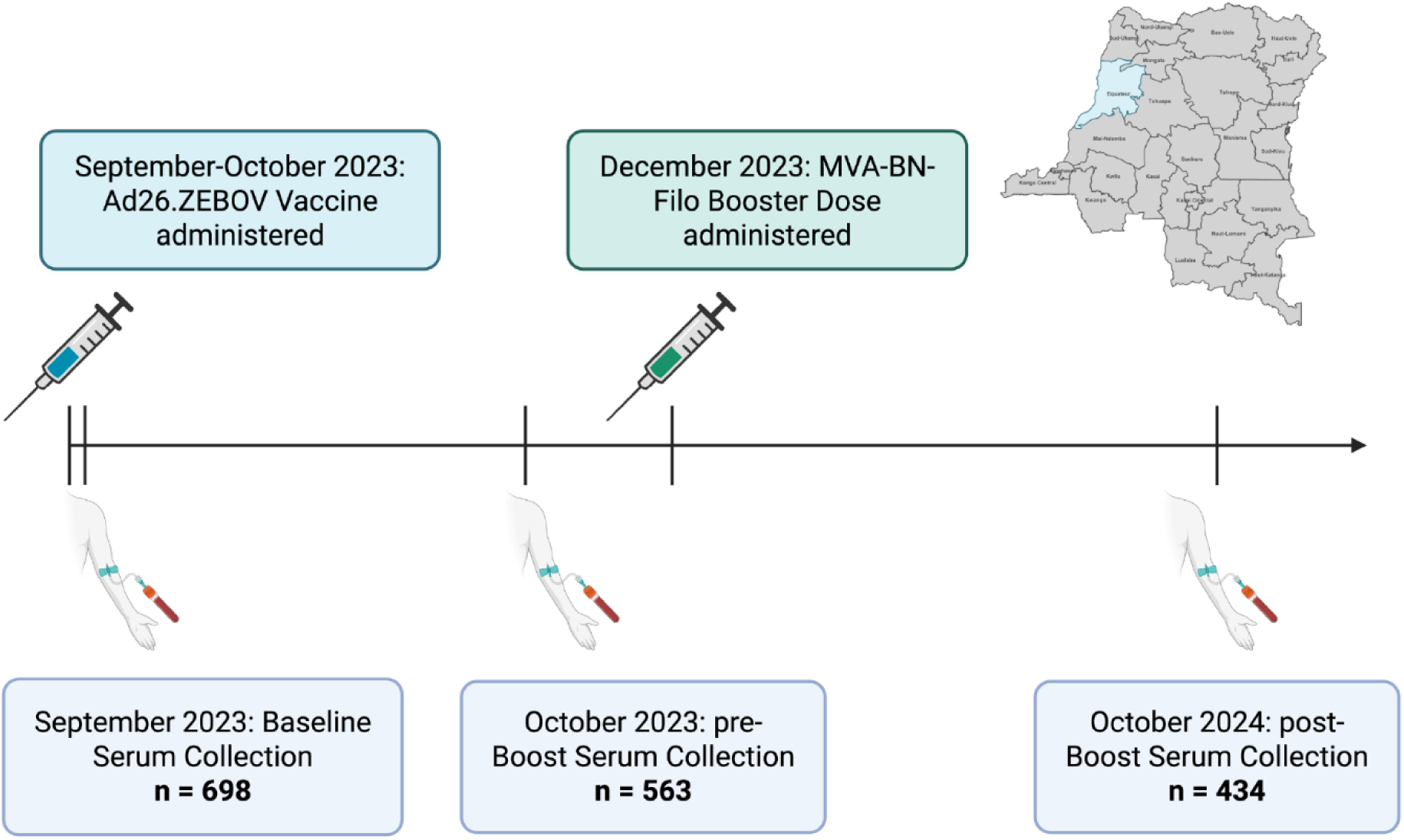
Study timeline and location of recruitment, Equateur province, DRC. Created using BioRender.com.

At baseline, 48·7% (*n* = 340, 95% CI: 45·0 - 52·4) of participants displayed seroreactivity against one or more of the six filovirus antigens, referred to hereafter as overall seroreactivity (**Figure 2A,B**, **Table 1**). The majority of these individuals displayed baseline seroreactivity against BDBV GP (31·1%, 95% CI: 27·8 - 34·6; **Table 1**). Seroreactivity against the other antigens varied, with 15·2% of individuals having reactivity against EBOV GP (95% CI: 12·7 - 18·0), 10·7% against EBOV VP40 (95% CI: 8·6 - 13·3), 8·9% against SUDV GP (95% CI: 7·0 - 11·2), 13·6% against MARV GP (95% CI: 11·3 - 16·4), and 5·4% against MARV VP40 (95 % CI: 4·0 - 7·4) (**Figure 2A-C**).

**Figure 2.**
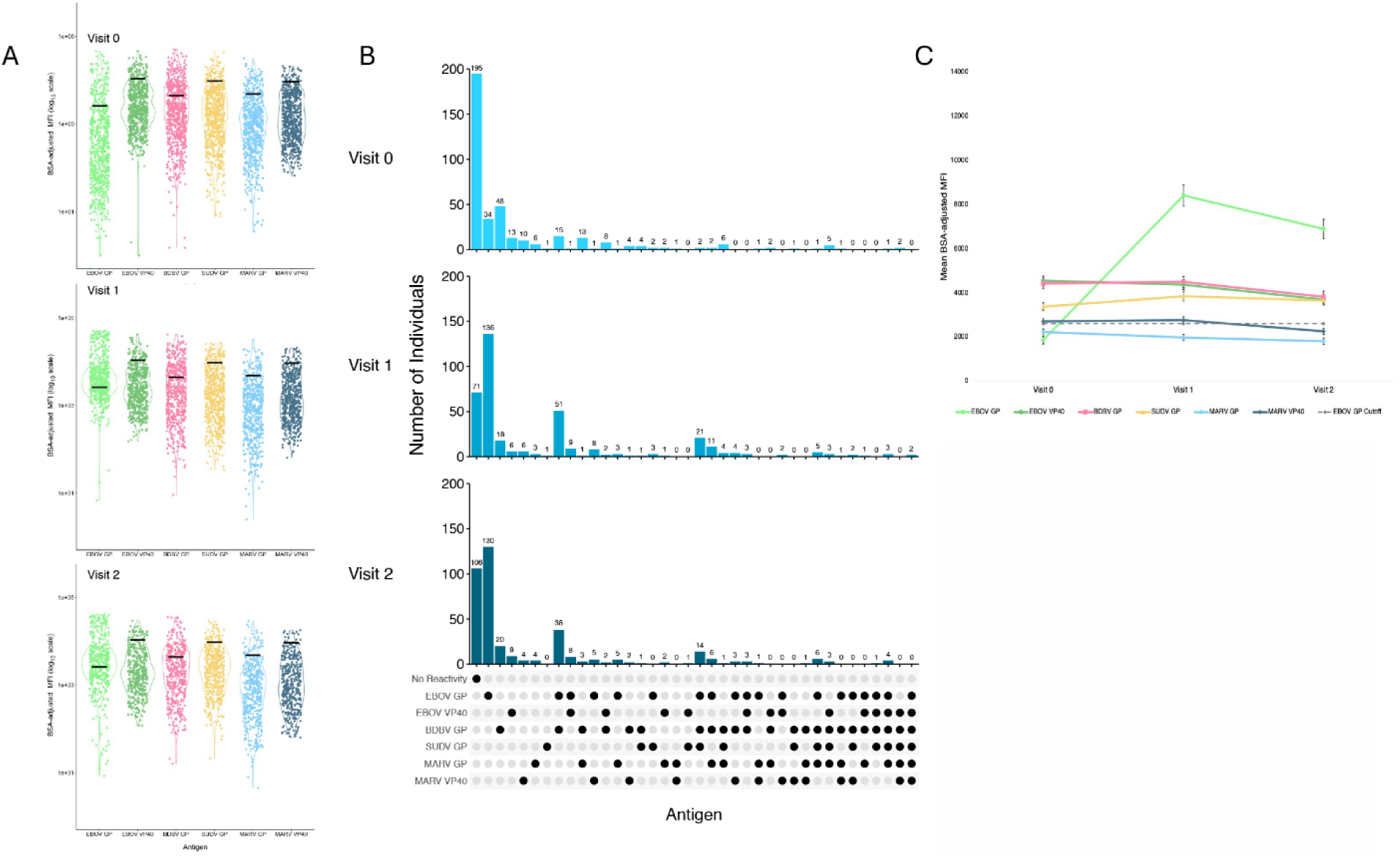
Cohort-wide reactivity against EBOV GP increased following vaccination and remained elevated one year later at visit 2. (**A/C**) Includes all participants at each visit: visit 0 (*n*=698), visit 1 (*n*=563), visit 2 (*n*=434). (**A**) Data are presented as BSA-adjusted mean fluorescence intensity (MFI) for reactivity to each of the antigens at each visit with standard error. Grey dashed line is the cutoff for reactivity against EBOV GP. (**B**) Upset plots showing seroreactivity among all individuals who attended all three visits (*n*=383) at each visit, stratified by number of individuals with seroreactivity to the specific antigen or combination of antigens. The plot is ordered from left to right by increasing seroreactivity to multiple antigens. (**C**) Violin plots presented as the log of BSA-adjusted MFI, black lines indicate the pre-determined threshold for reactivity for each individual antigen.

**Table 1.**
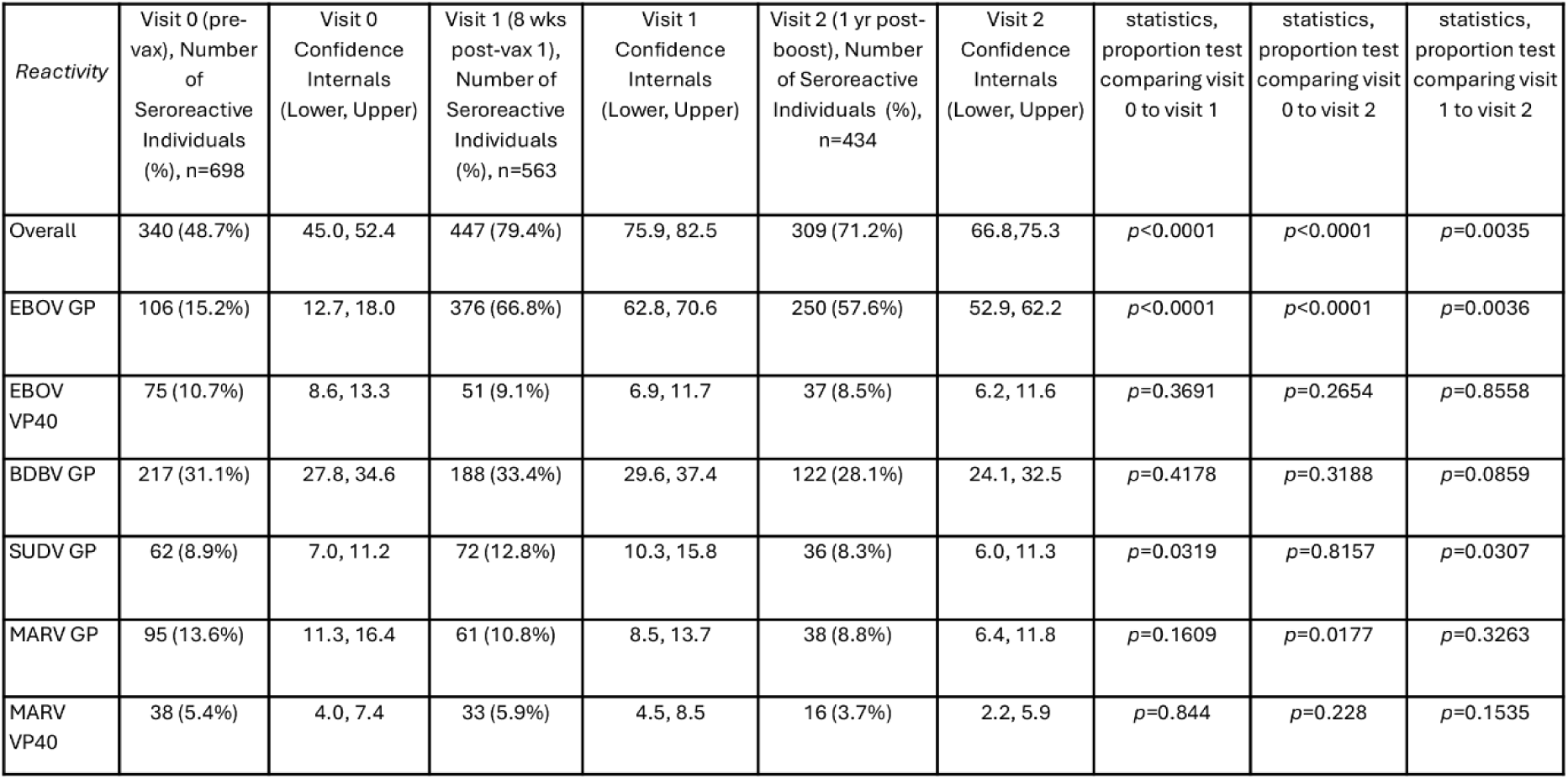
Overall seroreactivity by antigen at each visit for all participants, where overall reactivity refers to reactivity to any one or more of the antigens of interest.

### The frequency of reactivity against multiple antigens increased following vaccination

To assess the changes in frequency of antigen reactivity over the study period, we performed subgroup analyses on only the individuals who attended all three study visits (*n*=383). Among this group at baseline, the frequency of having no reactivity was much higher than the frequency of having seroreactivity to any of the filovirus antigens. This decreased substantially following vaccination, as would be expected, with the majority of individuals having reactivity to EBOV GP at visits 1 and 2. However, there were some individuals who never developed reactivity against EBOV GP despite vaccination (**Figure 2B**). Overall, there were increased numbers of individuals with reactivity to multiple antigens following vaccination, which may suggest a slight broadening of reactivity following vaccination, but this requires further confirmation as part of future studies.

Seroreactivity was significantly higher among male than female participants at baseline with 53·9% (95% CI: 49·1 - 58·6) of male participants and 40·7% (95% CI 34·8 - 46·9) of female participants having seroreactivity against one or more of the antigens of interest at visit 0 (*p*= 0·0012, **Supplementary Table 1**). Relative MFI of reactivity against each of the antigens was also significantly higher among males than females for all antigens (*p*<0.05; **Figure 3A**).

**Figure 3.**
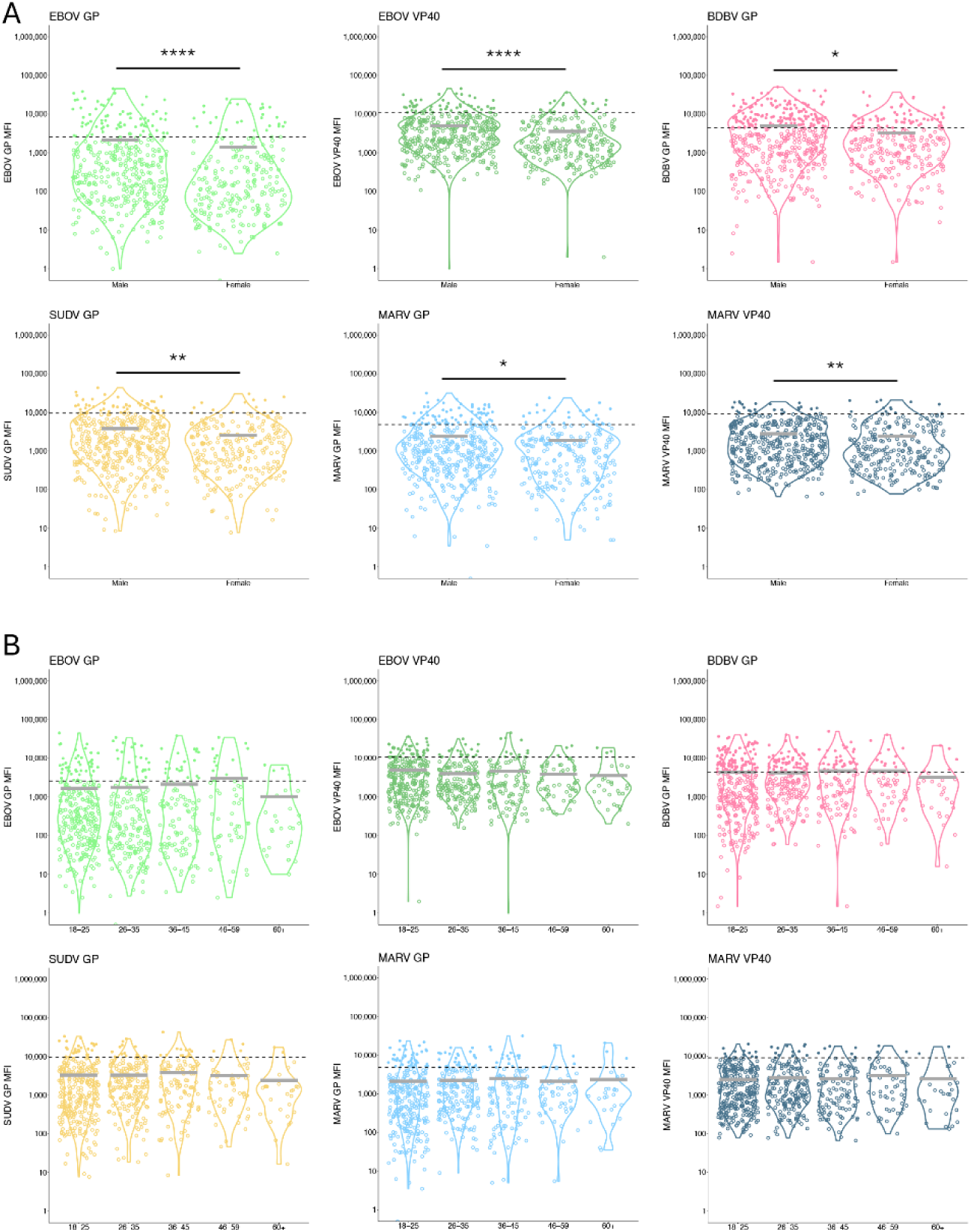
Baseline (visit 0) reactivity varies among the cohort by sex but not age. BSA-adjusted MFI for each antigen at visit 0 for all participants (n=698). Data presented as log of BSA-adjusted MFI, where the dashed black line denotes previously-established thresholds for seroreactivity for each antigen. Solid grey lines denotes mean MFI for each sub-group. Filled dots represent individuals who had reactivity above the cutoff value while the dots with only outlines represent individuals who do not pass the cutoff for seroreactivity. (**A**) Reactivity by antigen for males (*n*=421) and females (*n*=253). Mean MFI compared by ANOVA. **** p<0.0001, *** p<0.0005, **p<0.005, *p<0.05. (**B**) Comparison of reactivity for each age category, 18-25 (*n*=295 ), 26-35 (*n*=184), 36-45 (*n*=110), 46-59 (*n*=56), 60+ (*n*=27). Using pairwise Wilcox tests, none of the comparisons between age groups for any of the antigens were significant.

Reactivity at baseline among the various age groups was quite similar, with the highest reactivity being among those aged 36-45 (53·6%) and the lowest among those aged 60+ (37·0%, *p*>0·05; **Figure 3B**, **Supplementary Table 2**). When comparing the proportions of individuals with reactivity against each of the antigens for each age group, only two groups differed significantly when compared to the youngest group (18-25). The proportion of individuals with reactivity against MARV GP was significantly elevated among individuals aged 36-45 (14·3% [95% CI: 10·7 - 18·8] vs. 38·2% [95% CI: 29·6 - 47·5], *p*<0·0001) and the proportion of individuals with reactivity against MARV VP40 was significantly higher among individuals aged 46-59 (4·4% [95% CI: 2·5 - 7·5] vs. 5·4% [95% CI 1·3 - 15·2], *p*=0·038, **Supplementary Table 2**). MFI by antigen did not differ significantly between any of the age groups (**Figure 3B**).

The proportion of individuals with seroreactivity against any filovirus antigen at baseline was significantly higher among participants who identified as health care providers than those who did not (71·1% [95% CI: 55·1 - 83·1] vs 46·0% [95% CI: 42·0 - 49·7], *p*=0·0043). Baseline seroreactivity against EBOV GP was also significantly higher among participants who identified as health care providers than the rest of the participants (63·2% [95% CI: 47·2 - 76·7] vs 12·4% [95% CI: 10·1 - 15·2], *p*<0·0001; **Figure 4A**). MFI also varied significantly between health care providers and non-health care providers with higher mean MFI for EBOV GP reactivity reported among health care providers (8892 vs. 1466, *p*<0·0001; **Figure 4B**). Interestingly, although the proportions of individuals with reactivity against EBOV VP40 did not significantly differ between the two groups, the mean MFI of reactivity against EBOV VP40 was significantly higher among non-health care providers than health care providers (4645 vs 2088, *p*=0·0005; **Figure 4B**).

**Figure 4.**
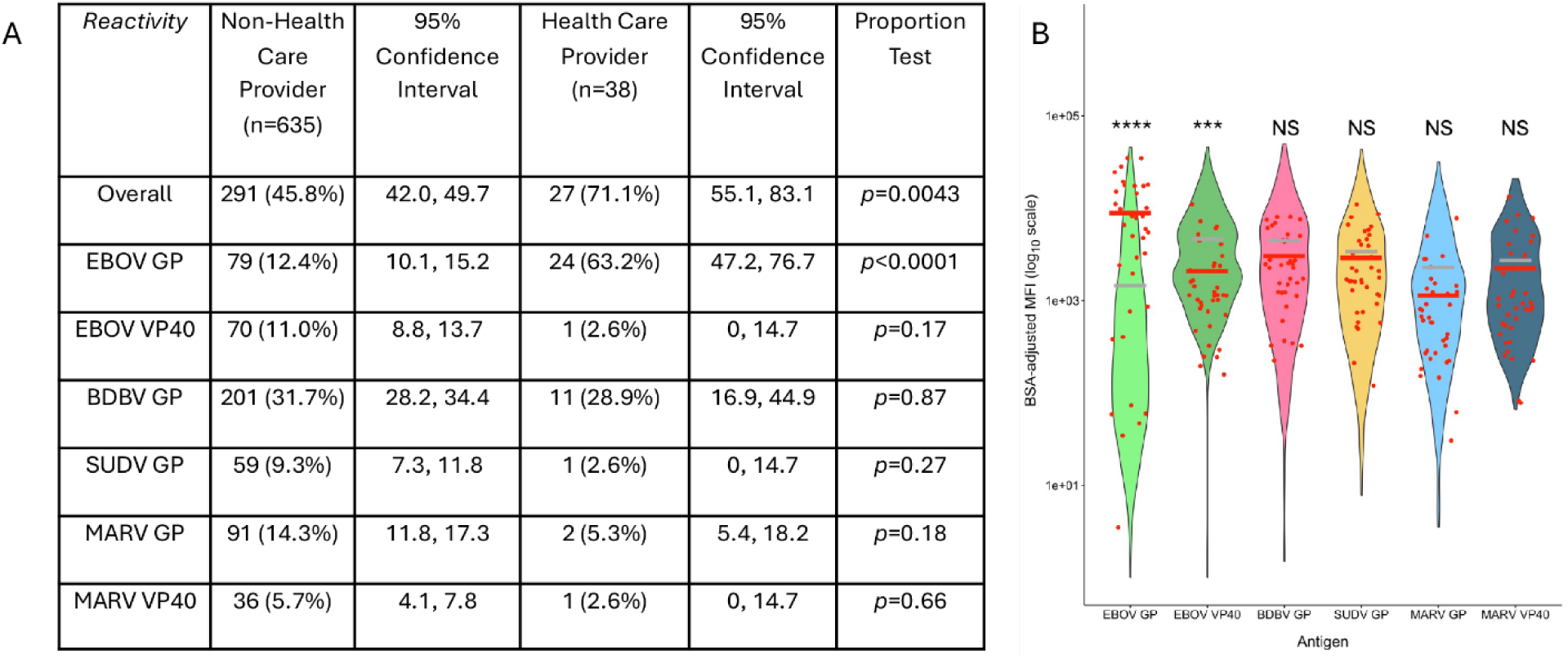
Baseline (visit 0) reactivity varies among the cohort by health care provider status. (**A**) Proportional reactivity stratified by health care provider status at visit 0, where overall reactivity refers to reactivity to any one or more of the antigens of interest. (**B**) BSA-adjusted MFI for each antigen at visit 0. Data presented as log MFI (**A**) Individuals who identified as health care providers are represented as red dots (*n*=38/698), on top of the distribution of the rest of the study population (*n*=635/698). Mean MFI of the health care providers is represented by the red bar. The grey line represents mean MFI of the rest of the population. Mean MFI was compared for each antigen by ANOVA. **** p<0.0001, *** p<0.0005.

### Reactivity against EBOV GP

As expected, the overall number of individuals with seroreactivity against EBOV GP among the entire cohort increased from 15·2% (95% CI: 12·7 - 18·0) at visit 0 to 66·8% (95% CI: 62·8 - 70·6) at visit 1, following the administration of the first dose of vaccine (*p*<0·0001) and to 57·6% (95% CI: 52·9 - 62·2, *p*<0·0001) at visit 2 (**Figure 5A**). At visit 2, compared to one year before at visit 1, EBOV GP seroreactivity decreased significantly among the overall cohort to 57·6% (95% CI: 52·9 - 62·2, *p*=0·0036).

**Figure 5.**
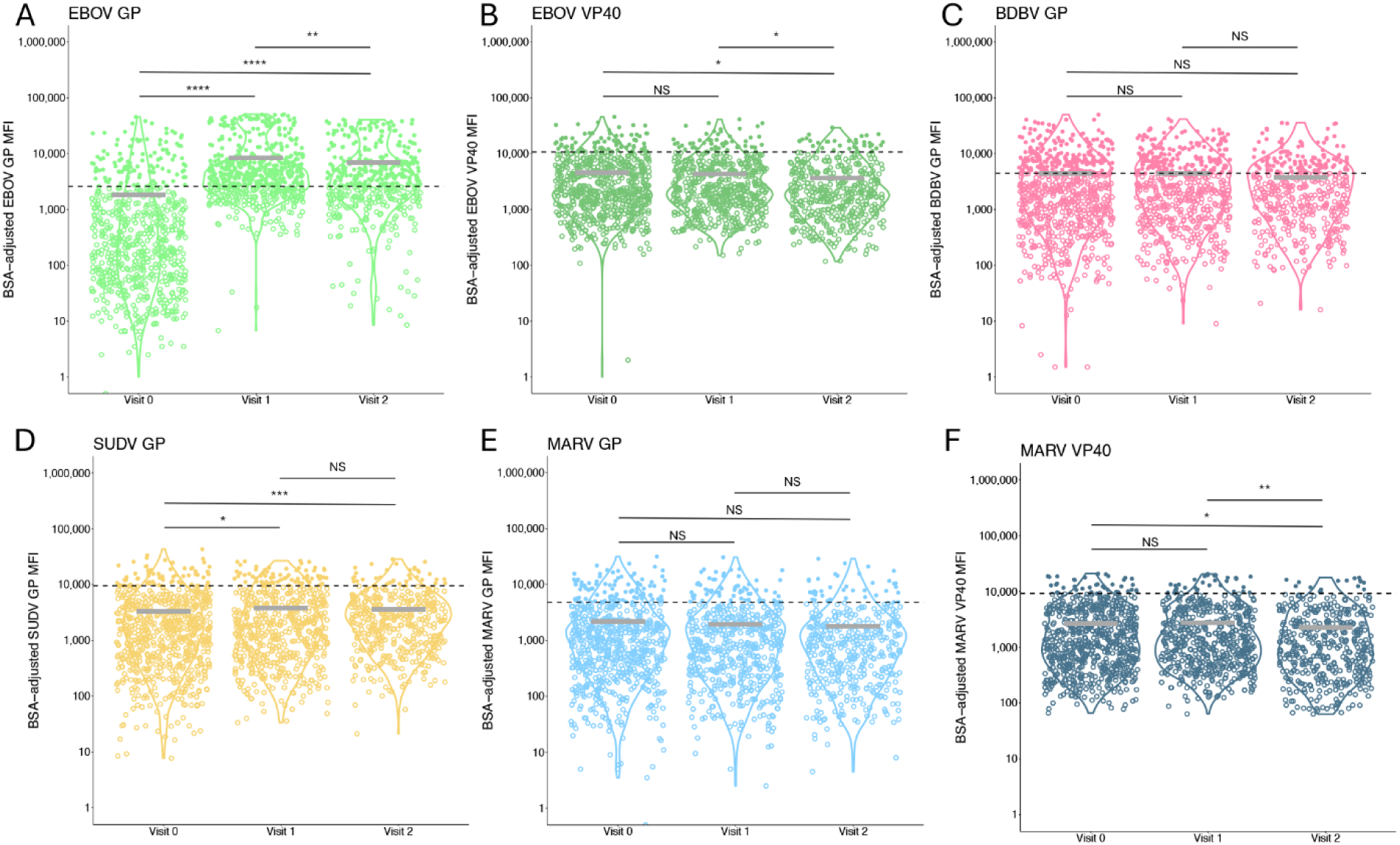
Reactivity changed over the course of the study. Log of BSA-adjusted MFI at three study visits for all participants at each visit, visit 0 (*n*=698), visit 1 (*n*=563), visit 2 (*n*=434). (**A**) EBOV GP, (**B**) EBOV VP40, (**C**) BDBV GP, (**D**) SUDV GP, (**E**) MARV GP, (**F**) MARV VP40. (**A-F**) Dashed black line denotes previously-established thresholds for seroreactivity. Solid grey lines denotes mean MFI for each antigen at each visit. Filled dots represent individuals who had reactivity above the cutoff value while the dots with only outlines represent individuals who do not pass the cutoff for seroreactivity. Variance compared using pairwise Wilcox test. **** p<0.0001, *** p<0.0005, **p<0.005, *p<0.05.

Among those participants considered naïve at baseline, EBOV GP reactivity increased significantly at visit 1 to 61·5% (95% CI: 55·8 - 67·0, *p*<0·0001) but decreased to 50·9% (95% CI: 44·3 - 57·6, *p*<0·05) at visit 2, one year post-vaccination. Similar to the entire cohort, the proportion of individuals with reactivity to EBOV GP decreased significantly between visit 1 and visit 2 (*p*=0·023; **Figure 5A**).

### Reactivity against other filovirus antigens

Cohort-wide, the proportion of individuals with reactivity against EBOV VP40 did not change throughout the study duration (*p*>0·05 for all timepoints, **Figure 5B**, **Table 1**). However, when looking at either the naive population or the baseline reactive population, there were significant changes in the proportion of individuals with EBOV VP40 reactivity when comparing visit 0 to visit 2. For those that were naïve at baseline, proportional reactivity increased from 0% (95% CI 0·0 - 0·013) to 2·8% (95% CI: 1·2 - 6·1; *p*=0·0058, **Supplementary Figure 1**) and those that were reactive at baseline decreased significantly from 22·0% (95% CI: 17·9 - 26·7) to 14·4% (95% CI: 10·3 - 19·9; *p*=0·038; **Supplementary Figure 2**).

The proportion of individuals with anti-BDBV antibodies did not significantly change over the study period among the entire cohort (*p*>0·05; **Figure 5C**, **Table 1**). However, among naïve individuals, BDBV GP reactivity increased significantly between visits 0 (0% [95% CI: 0·0 - 0·013]) and 1 (5·6% [95% CI: 3·4 - 9·0]; *p*<0·0001) and visits 0 and 2 (9·3% [95% CI: 6·1 - 14·1], *p*<0·0001; **Supplementary Figure 1**). To aid in understanding potential baseline cross-reactivity against BDBV, we also looked at reactivity among those who did not display anti-EBOV reactivity at baseline to remove any individuals who had received either of the EBOV vaccines previously, even though prior vaccination was an exclusion criterion for the study. Following removal of EBOV GP seroreactivity, many individuals still displayed reactivity against the other filovirus antigen, with 28·9% (95% CI: 25·3 - 32·6) of individuals having reactivity against BDBV GP (**Supplementary Figure 3**).

Reactivity to SUDV GP among all participants increased from 8·9% (95% CI: 7·0 - 11·2) to 12·8% (95% CI: 10·3 - 15·8) at visit 1 before returning to 8·3% (95% CI: 6·0 - 11·3) at visit 2 (**Figure 5D**, **Table 1**). Among those naïve at baseline there were no significant increases or decreases in the proportion of individuals with SUDV GP reactivity observed (**Supplementary Figure 1**). Additionally, MARV GP responses decreased as the study progressed, with a significant decrease between visits 0 and 2 (13·6% [95% CI: 11·3 - 16·4] vs 8·8% [95% CI: 6·4 - 11·8], *p*=0·018, **Figure 4E**, **Table 1**). However, among those who were naive at baseline, there was a significant increase in the proportion of individuals with reactivity against MARV GP between visit 0 and visit 2 (0% [95% CI: 0·0 - 0·013] vs. 2·8% [95% CI: 1·2 - 6·1], *p*=0·0058; **Supplementary Figure 1**).

The proportion of individuals with reactivity against MARV VP40 among the entire cohort did not change significantly throughout the study period (*p*>0·05 at all visits, **Figure 5F**, **Table 1**). However, among those individuals who were naïve at baseline, the number of individuals with reactivity increased significantly from visit 0 (0% [95% CI: 0·0 - 0·013]) to visit 1 (2·4% [95% CI: 1·1 - 5·1], *p*=0·0096) and from visit 0 to visit 2 (1·9% [95% CI: 0·6 - 4·9], *p*=0·038; **Supplementary Figure 1**).

## Discussion

Here, we assessed Ad26.ZEBOV-MVA-BN-filo vaccination immunogenicity in, to our knowledge, the largest cohort of vaccinated individuals in a region endemic for multiple filoviruses that cause disease in humans. The deployment of Ad26.ZEBOV-MVA-BN-filo vaccine among this population was outside of an outbreak context and was aimed at increasing population-level protection against EVD. Our investigation showed high levels of baseline seroreactivity against multiple filoviruses among individuals recruited from EQT Province, DRC, an area that has had several previous outbreaks of EVD since 2018, with the most recent outbreak in 2022^3^, and EBOV GP seroreactivity increased substantially following administration of the Ad26.ZEBOV-MVA-BN-filo vaccine.

Baseline seroreactivity, particularly against BDBV GP, was elevated at baseline and remained elevated throughout the study. However, there have been no documented outbreaks of BDBV in this area of DRC. Recent results from our team have documented elevated seroreactivity to BDBV GP among a different cohort in the same region between 2021 and 2022, corresponding to 2.5-3.5 years post-rVSVΔG-ZEBOV-GP (Ervebo) vaccination.^20^ Baseline seroreactivities against non-EBOV filovirus GPs is likely not attributable to previous Ebola vaccination, as prior vaccination was an exclusion criteria. Further, we detected reactivity against the other filovirus antigens even after individuals with EBOV GP reactivity were removed from analyses.

Baseline seroreactivity varied substantially by sex for nearly all antigens, though only by health care provider status for EBOV GP. These differences in baseline reactivity by sex (possibly indirectly linked to traditional gender roles) and whether the individual identifies as a health care provider were expected, and are known risk factors for filovirus infection.^21,22^ Seroreactivity did not vary significantly among the cohort by age grouping, with the exception of the reactivity to EBOV GP post-vaccination, although this was expected, as younger individuals tend to respond more strongly to vaccinations than older adults (reviewed by Fink *et al*^23^).

Seroreactivity against EBOV GP increased significantly among the cohort following vaccination with the Ad26.ZEBOV-MVA-BN-filo vaccine, at eight weeks post-primary vaccination, 66.8% of individuals had seroreactivity against EBOV GP. This varies significantly from previous studies which documented seroconversion among all study participants.^24^ At one year post-booster dose of vaccine, anti-EBOV GP reactivity remained elevated compared to baseline, with 57.6% of individuals displaying seroreactivity against EBOV GP, suggesting the vaccine induced relatively durable and stable immune responses to EBOV GP among those who responded. Other studies have evaluated EBOV GP reactivity rates following vaccination with rVSVΔG-ZEBOV-GP using the same assay and have noted similar peaks of reactivity but less waning at one-year post-vaccination^25^ compared to what we report here.

While some efficacy studies of the Ad26.ZEBOV-MVA-BN-Filo are currently lacking, Mdluli *et al*. reported on the immunogenicity of this heterologous vaccine in a study spanning six countries, showing induction of high levels of antibody binding against EBOV GP, antibodies produced with Fc effector functions, and neutralizing antibodies in conjunction with functional CD4+ T cell responses.^26^ Similar to our findings, despite the inclusion of the GPs from SUDV and MARV in the booster dose, Mdluli *et al.* reported only a slight increase in antibody responses against SUDV and no change in anti-MARV GP antibodies. As the peak prevalence of SUDV reactivity in the current study was documented before the boost dose given after visit 1, this suggests there was no evidence of increased antibody response following booster administration. Together, these data suggest that this vaccine induces minimal antibody responses against other filoviruses and may, therefore, have minimal benefit in a non-EBOV filovirus outbreak.

To date, the efficacy and immune responses induced by the rVSVΔG-ZEBOV-GP vaccine have been more widely studied and well-characterized than the Ad26.ZEBOV-MVA-BN-Filo vaccine. Protection by rVSVΔG-ZEBOV-GP against lethal EBOV infection of non-human primates was shown to be dependent on the presence of antibodies against the virus GP.^16,27^ However, neutralizing antibodies did not seem to be induced at high levels despite the vaccine being protective, suggesting that non-neutralizing antibodies may play a role in protection and that their presence may be predictive of protection.^27^ Cross-protection induced by rVSVΔG-ZEBOV-GP has been investigated in laboratory settings with discordant conclusions. While some reports suggest rVSVΔG-ZEBOV-GP induced cross-protection against or cross-reactive antibodies against BDBV^13,17,28^, other studies reported no protection against BDBV disease and death following rVSVΔG-ZEBOV-GP vaccination.^29^ Correlates of protection have not yet been established for the Ad26.ZEBOV-MVA-BN-Filo vaccine used here or for BDBV survival in humans, but it is likely that the immunological correlates of protection are similar to those for rVSVΔG-ZEBOV-GP, pointing to important roles for non-neutralizing antibodies as well as roles for other immune cells.

### Limitations

While non-human primate studies have suggested that non-neutralizing antibodies are needed for protection against EVD, the correlates of protections in humans remain unclear.^16,27^ Therefore, we could not determine if seroreactivity detected using this antigen-based assay indicates real-world protection against EVD in this cohort, although previous studies have reported induction of anti-EBOV GP neutralizing antibodies in response to this vaccine.^26^ The MIA used detects antibody responses to selected filovirus antigens and thus does not allow further identification or characterization of the antibodies, so we are unable to make conclusions regarding functionality of the antibodies detected (whether neutralizing, etc). Additionally, loss to follow up during the study period, could have led to response biases. However, this study did have a 62% (*n*=434/698) retention rate, which is considered high and is in alignment with other recent studies in the DRC.^25^ Other limitations of the study include the lack of 8-week post-boost follow-up as an additional time point to detect immediate changes in reactivity as well as unknown underlying medical status, such as infections with HIV or other immunosuppressive conditions that could influence vaccine-induced immune responses. However, well-controlled HIV among those on anti-retroviral therapies does not seem to affect immune responses to this vaccine.^30^

Future studies would benefit from investigations into antibody responses against other filovirus proteins, particularly those of BDBV to further confirm the extent of previous exposures to BDBV within this or other cohorts in the DRC. Understanding the extent of previous exposures and pre-existing immunity to BDBV would certainly have real-world implications for the ongoing and future BDBV outbreaks. Among those individuals with reactivity against BDBV, it would be particularly interesting to determine whether these individuals displayed reactivity against other BDBV proteins and whether the antibodies detected as part of the current study are neutralizing. Additional laboratory analyses are also needed to determine whether neutralizing antibodies are induced, particularly those with cross-protective effects induced by the Ad26.ZEBOV-MVA-BN-Filo vaccine. As this report is not the first to show minimal antibody responses induced by the other antigens included in the booster dose of this vaccine, improvements have already been suggested with considerable interest in inducing a stronger immune response. Recent work utilized a next-generation vaccinia vector (Sementis Copenhagen vector) expressing the EBOV GP, which when used as a boost following Ad26.ZEBOV as the first vaccine, induced a robust T cell response and antibody titres similar to those reported previously in non-human primates. However, the efficacy of this vaccine was not assessed and only EBOV GP was included as an antigen.^31^

## Conclusions

Taken together, the results presented here report substantial baseline reactivity against filoviruses in EQT province of DRC, particularly to BDBV GP, raising the question as to whether previous BDBV outbreaks went unidentified. Additionally, we report significant increases in reactivity to EBOV GP following vaccination with Ad26.ZEBOV-MVA-BN-Filo that was maintained for at least one year-post vaccination, with minimal changes to reactivity against other filovirus antigens post-vaccination.

## Data Availability

All data produced in the present study are available upon reasonable request to the authors.

## Acknowledgements

The authors thank all study participants and Dr. Jordan Wight and Ms. Morgan Hiebert for their consultation regarding data analysis and visualization. The authors would also like to thank the local phlebotomists, interviewer staff and participants in Mbandaka, EQT, without whom this work would not be possible.

## Author contributions

Conceptualization: AWR, NAH, PMK, PM, LEH

Data Curation: HLW, NAH

Formal Analysis: HLW, MH

Funding Acquisition: JK, AWR, NAH, LEH, PMK, PKM

Investigation: HLW, SM, NAH, JPK, MB, HK, MT, ZM, HK

Methodology: HLW, AHB, AT, TW, ATL, AWR, NAH, MH

Project Administration: NAH, AWR, PMK, PM

Resources: AWR, PMK. ATL

Software: HLW

Supervision: NAH, PKM, PMK, JK, AWR

Validation: HLW

Visualization: HLW, MH, SM

Writing - original draft: HLW

Writing - review and editing: all authors

## Supplementary Data

**Supplementary Figure 1.**
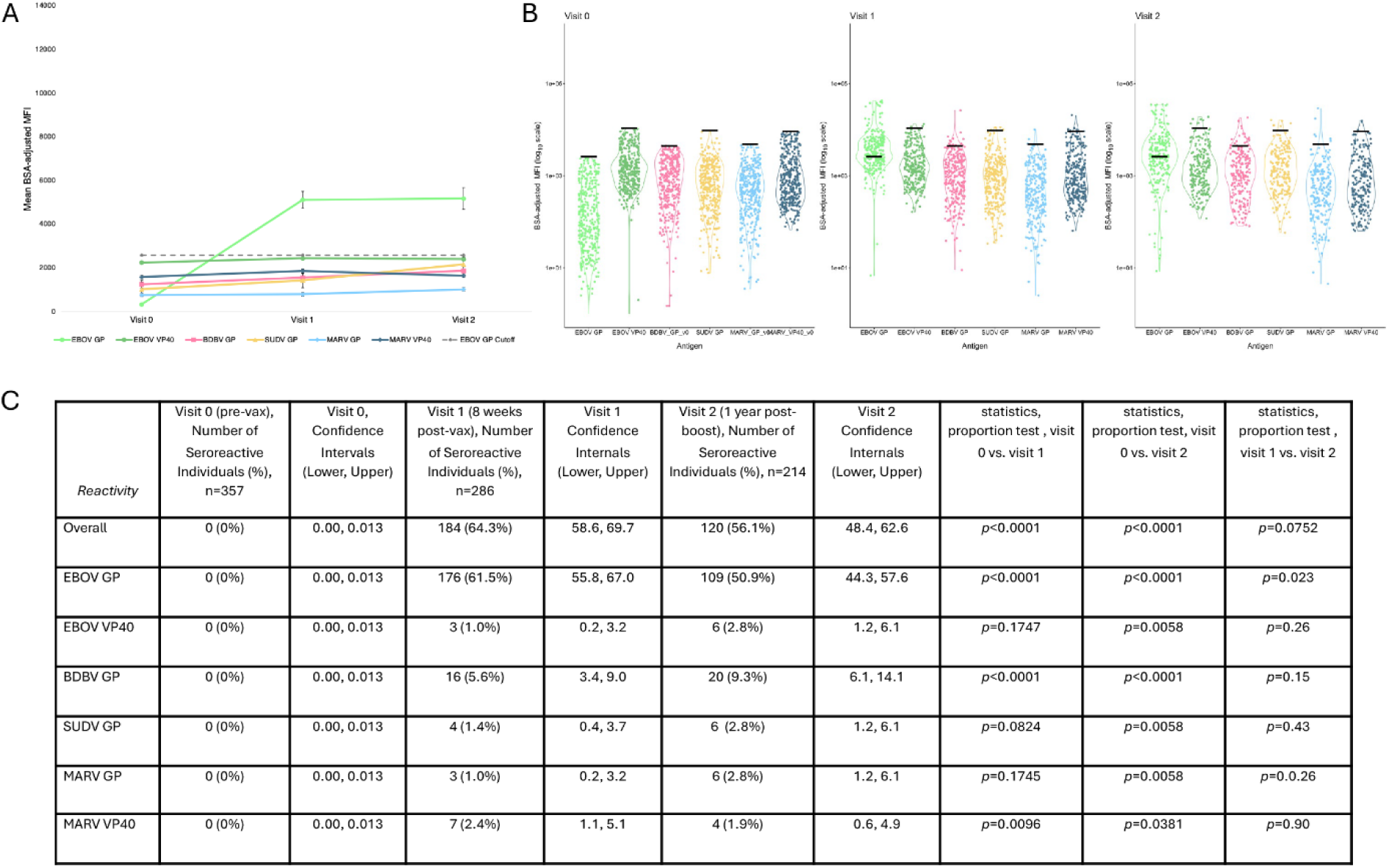
Reactivity among those who were naïve at baseline. **(A-C)** Only participants that had no reactivity at visit 0. Visit 0 (*n*=357), visit 1 (*n*=286), visit 2 (*n*=214). (**A**) Data are presented as BSA-adjusted MFI for reactivity to each of the antigens at each visit with standard error. Grey dashed line is the cutoff for reactivity against EBOV GP. (**B**) Violin plots presented as the log of BSA-adjusted MFI at each timepoint, black lines indicate the pre-determined threshold for reactivity for each individual antigen. (**C**) Seroreactivity by antigen at each visit, those with any baseline reactivity removed from analysis, where overall reactivity refers to reactivity to any one or more of the antigens of interest.

**Supplementary Figure 2.**
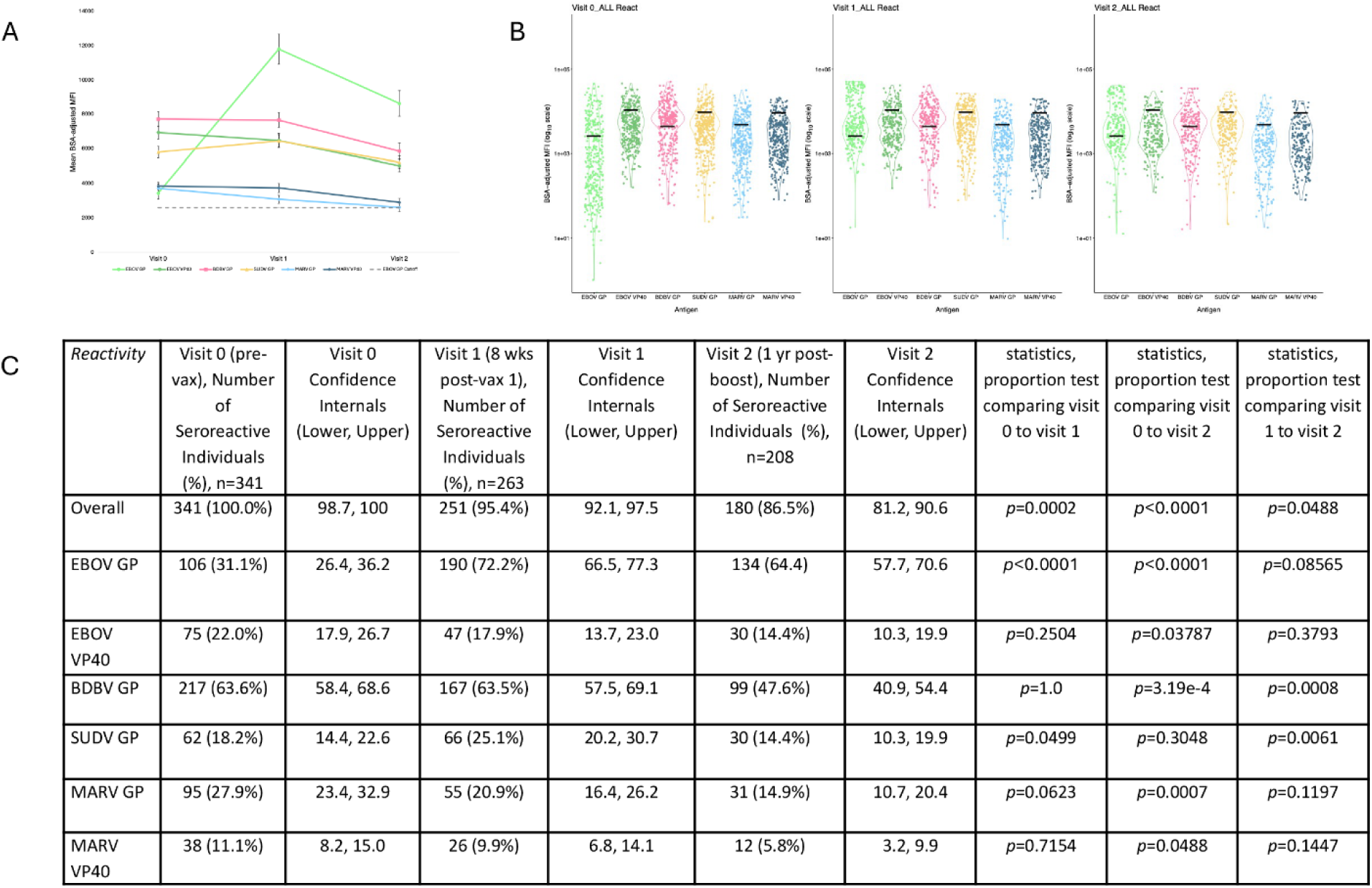
Reactivity among those who displayed reactivity at baseline. (**A-C**) Only participants that had reactivity at visit 0. Visit 0 (*n*=341), visit 1 (*n*=263), visit 2 (*n*=208). (**A**) Data are presented as BSA-adjusted MFI for reactivity to each of the antigens at each visit with standard error. Grey dashed line is the cutoff for reactivity against EBOV GP. (**B**) Violin plots presented as the log of BSA-adjusted MFI at each timepoint, black lines indicate the pre-determined threshold for reactivity for each individual antigen. (**C**) Overall seroreactivity by antigen at each visit for only those that displayed seroreactivity at baseline, where overall reactivity refers to reactivity to any one or more of the antigens of interest.

**Supplementary Figure 3.**
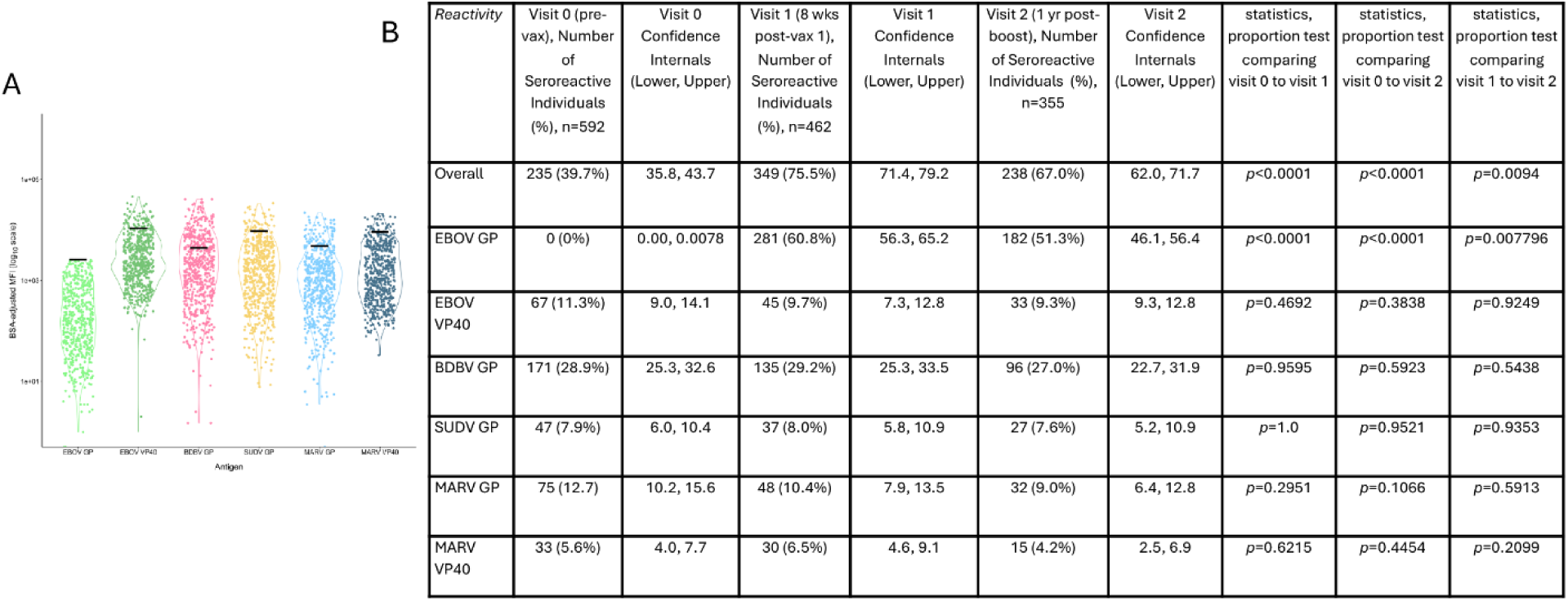
BDBV reactivity is not dependent on EBOV GP reactivity. (**A**) Log of BSA-adjusted MFI for each antigen at visit 0 with those who displayed EBOV GP reactivity removed (leaving *n*=592). Black line denotes seroreactivity cutoffs for each antigen. (**B**) Overall seroreactivity by antigen at each visit with those with baseline reactivity against EBOV GP removed, where overall reactivity refers to reactivity to any one or more of the antigens of interest.

**Supplementary Table 1.** Overall seroreactivity by antigen at each visit, stratified by sex, where overall reactivity refers to reactivity to any one or more of the antigens of interest.

| Reactivity | Number of Seroreactive Males | Confidence Internals, Males (Lower, Upper) | Number of Seroreactive Females | Confidence Internals, Females (Lower, Upper) | statistics, proportion test comparing Males vs Females |
| --- | --- | --- | --- | --- | --- |
| Overall |  |  |  |  |  |
| Visit 0 | 227/421 (53.9%) | 49.1, 58.6 | 103/253 (40.7%) | 34.8, 46.9 | $p=0.00118$ |
| Visit 1 | 268/335 (80%) | 75.4, 83.9 | 179/228 (78.5%) | 72.7, 83.4 | $p=0.7465$ |
| Visit 2 | 186/254 (73.2%) | 67.5, 78.3 | 119/174 (68.4%) | 61.1, 74.8 | $p=0.3283$ |
| EBOV GP |  |  |  |  |  |
| Visit 0 | 75/421 (17.8%) | 14.5, 21.8 | 28/253 (11.1%) | 7.7, 15.6 | $p=0.02465$ |
| Visit 1 | 216/335 (64.5%) | 59.2, 69.4 | 160/228 (70.2%) | 63.9, 75.7 | $p=0.1875$ |
| Visit 2 | 150/254 (59.1%) | 52.9, 64.9 | 96/174 (55.2%) | 47.7, 62.4 | $p=0.4848$ |
| EBOV VP40 |  |  |  |  |  |
| Visit 0 | 51/421 (12.1%) | 9.1, 15.4 | 21/253 (8.3%) | 5.4, 12.4 | $p=0.1547$ |
| Visit 1 | 36/335 (10.7%) | 7.8, 14.5 | 15/228 (6.6%) | 4.0, 10.6 | $p=0.1232$ |
| Visit 2 | 27/254 (10.6%) | 7.4, 15.1 | 10/174 (5.7%) | 3.0, 10.4 | $p=0.1117$ |
| BDBV GP |  |  |  |  |  |
| Visit 0 | 153/421 (36.3%) | 31.7, 40.9 | 60/253 (23.7%) | 18.9, 29.3 | $p=0.0008792$ |
| Visit 1 | 129/335 (38.5%) | 33.5, 43.8 | 59/228 (25.9%) | 20.6, 31.9 | $p=0.002459$ |
| Visit 2 | 81/254 (31.9%) | 26.5, 37.9 | 39/174 (22.4%) | 16.8, 29.2 | $p=0.04193$ |
| SUDV GP |  |  |  |  |  |
| Visit 0 | 47/421 (11.2%) | 8.5, 14.6 | 13/253 (5.1%) | 2.9, 8.7 | $p=0.01173$ |
| Visit 1 | 53/335 (15.8%) | 12.3, 20.1 | 19/228 (8.3%) | 5.3, 12.7 | $p=0.01303$ |
| Visit 2 | 25/254 (9.8%) | 6.7, 14.2 | 10/174 (5.7%) | 3.0, 10.4 | $p=0.1805$ |
| MARV GP |  |  |  |  |  |
| Visit 0 | 68/421 (16.2%) | 13.0, 20.0 | 25/253 (9.9%) | 6.7, 14.2 | $p=0.02998$ |
| Visit 1 | 40/335 (11.9%) | 8.9, 15.9 | 21/228 (9.2%) | 6.0, 13.7 | $p=0.3762$ |
| Visit 2 | 25/254 (9.8%) | 6.7, 14.2 | 13/174 (7.5%) | 4.3, 12.5 | $p=0.5002$ |
| MARV VP40 |  |  |  |  |  |
| Visit 0 | 25/421 (5.9%) | 4.0, 8.7 | 12/253 (4.7%) | 2.7, 8.2 | $p=0.6277$ |
| Visit 1 | 22/335 (6.6%) | 4.3, 9.8 | 11/228 (4.8%) | 2.6, 8.5 | $p=0.4957$ |
| Visit 2 | 8/254 (3.1%) | 1.5, 6.2 | 8/174 (4.6%) | 2.2, 9.0 | $p=0.6056$ |

**Supplementary Table 2.** Proportional reactivity against each antigen, stratified by age at each visit, where overall reactivity refers to reactivity to any one or more of the antigens of interest.

| Reactivity | 18 - 25 | 95%<br>Confidence<br>Interval | 26 - 35 | 95% Confidence<br>Interval | Proportion Test<br>(26-35<br>compared to 18-25) | 36 - 45 | 95%<br>Confidence<br>Interval | Proportion<br>Test (36-45<br>compared to<br>18-25) | 46 - 59 | 95%<br>Confidence<br>Interval | Proportion Test<br>(46-59<br>compared to<br>18-25) | 60+ | 95%<br>Confidence<br>Interval | Proportion Test<br>(60+ compared<br>to 18-25) |
| --- | --- | --- | --- | --- | --- | --- | --- | --- | --- | --- | --- | --- | --- | --- |
| Overall |  |  |  |  |  |  |  |  |  |  |  |  |  |  |
| Visit 0 | 141/295<br>(47.8%) | 42.2, 53.5 | 94/184<br>(51.1%) | 43.9, 58.2 | $p=0.5441$ | 59/110<br>(53.6%) | 44.4, 62.7 | $p=0.3504$ | 26/56<br>(46.4%) | 34.0, 59.3 | $p=0.9665$ | 10/27<br>(37.0%) | 21.5, 55.8 | $p=0.3838$ |
| Visit 1 | 200/240<br>(83.3%) | 78.1, 87.5 | 123/158<br>(77.8%) | 70.7, 83.7 | $p=0.2157$ | 72/95<br>(75.8%) | 66.2, 83.3 | $p=0.1506$ | 35/45<br>(77.8%) | 63.5, 87.6 | $p=0.493$ | 15/23<br>(65.2%) | 44.8, 81.3 | $p=0.0620$ |
| Visit 2 | 128/172<br>(74.4%) | 67.4, 80.4 | 86/123<br>(69.9%) | 61.3, 77.3 | $p=0.4706$ | 54/78<br>(69.2%) | 58.2, 78.4 | $p=0.4835$ | 28/38<br>(73.7%) | 57.8, 85.2 | $p=1.0$ | 8/16<br>(50.0%) | 28.0, 72.0 | $p=0.07243$ |
| EBOV GP |  |  |  |  |  |  |  |  |  |  |  |  |  |  |
| Visit 0 | 42/295<br>(14.2%) | 10.7, 18.7 | 24/184<br>(13.0%) | 8.9, 18.7 | $p=0.8162$ | 21/110<br>(19.1%) | 12.8, 27.5 | $p=0.2962$ | 12/56<br>(21.4%) | 12.6, 34.0 | $p=0.2439$ | 4/27<br>(14.8%) | 5.3, 33.1 | $p=1.0$ |
| Visit 1 | 180/240<br>(75.0%) | 69.1, 80.1 | 99/158<br>(62.7%) | 54.9, 69.8 | $p=0.0118$ | 57/95<br>(60.0%) | 50.0, 69.3 | $p=0.009683$ | 29/45<br>(64.4%) | 49.8, 76.8 | $p=0.1985$ | 9/23<br>(39.1%) | 22.1, 59.3 | $p=0.0006454$ |
| Visit 2 | 104/172<br>(60.5%) | 53.0, 67.5 | 68/123<br>(55.3%) | 46.5, 63.8 | $p=0.4413$ | 44/78<br>(56.4%) | 45.4, 66.9 | $p=0.6416$ | 24/38<br>(63.2%) | 47.2, 76.7 | $p=0.9011$ | 5/16<br>(31.3%) | 13.9, 55.9 | $p=0.04552$ |
| EBOV VP40 |  |  |  |  |  |  |  |  |  |  |  |  |  |  |
| Visit 0 | 36/295<br>(12.2%) | 8.9, 16.5 | 19/184<br>(10.3%) | 6.6, 15.6 | $p=0.6316$ | 10/110<br>(9.1%) | 4.8, 16.1 | $p=0.4827$ | 3/56<br>(5.4%) | 1.3, 15.2 | $p=0.2067$ | 3/27<br>(11.1%) | 3.0, 28.9 | $p=1.0$ |
| Visit 1 | 26/240<br>(10.8%) | 7.5, 15.4 | 12/158<br>(7.6%) | 4.3, 12.9 | $p=0.3674$ | 6/95<br>(6.3%) | 2.7, 13.4 | $p=0.2884$ | 3/45<br>(6.7%) | 1.6, 18.5 | $p=0.5621$ | 4/23<br>(17.4%) | 6.4, 37.7 | $p=0.5473$ |
| Visit 2 | 20/172<br>(11.6%) | 7.6, 17.3 | 7/123<br>(5.7%) | 2.6, 11.5 | $p=0.1239$ | 6/78<br>(7.7%) | 3.3, 16.1 | $p=0.471$ | 3/38<br>(7.9%) | 2.0, 21.5 | $p=0.704$ | 1/16<br>(6.3%) | 0.0, 30.3 | $p=0.8116$ |
| BDBV GP |  |  |  |  |  |  |  |  |  |  |  |  |  |  |
| Visit 0 | 91/295<br>(30.8%) | 25.8, 36.3 | 57/184<br>(31.0%) | 24.7, 38.0 | $p=1.0$ | 42/110<br>(38.2%) | 29.6, 47.5 | $p=0.2009$ | 17/56<br>(30.4%) | 19.8, 43.4 | $p=1.0$ | 5/27<br>(18.5%) | 7.7, 37.2 | $p=0.2624$ |
| Visit 1 | 69/240<br>(28.8%) | 23.4, 34.8 | 57/158<br>(36.1%) | 29.0, 43.8 | $p=0.1535$ | 37/95<br>(38.9%) | 29.7, 49.0 | $p=0.0932$ | 15/45<br>(33.3%) | 21.3, 48.0 | $p=0.6594$ | 10/23<br>(43.5%) | 25.6, 63.2 | $p=0.2173$ |
| Visit 2 | 46/172<br>(26.7%) | 20.7, 33.8 | 33/123<br>(26.8%) | 19.8, 35.3 | $p=1.0$ | 24/78<br>(30.8%) | 21.6, 41.8 | $p=0.6138$ | 12/38<br>(31.6%) | 19.0, 47.5 | $p=0.6871$ | 5/16<br>(31.3%) | 13.9, 55.9 | $p=0.9253$ |
| SUDV GP |  |  |  |  |  |  |  |  |  |  |  |  |  |  |
| Visit 0 | 27/295<br>(9.2%) | 6.3, 13.0 | 14/184<br>(7.6%) | 4.5, 12.5 | $p=0.6748$ | 13/110<br>(11.8%) | 6.9, 19.3 | $p=0.5402$ | 4/56<br>(7.1%) | 2.3, 17.5 | $p=0.8188$ | 2/27<br>(7.4%) | 1.0, 24.5 | $p=1.0$ |
| Visit 1 | 25/240<br>(10.4%) | 7.1, 15.0 | 19/158<br>(12.0%) | 7.8, 18.1 | $p=0.7358$ | 16/95<br>(16.8%) | 10.5, 25.7 | $p=0.152$ | 8/45<br>(17.8%) | 9.0, 31.6 | $p=0.2451$ | 4/23<br>(17.4%) | 6.4, 37.7 | $p=0.5018$ |
| Visit 2 | 12/172<br>(7.0%) | 3.9, 11.9 | 10/123<br>(7.3%) | 3.7, 13.5 | $p=0.8831$ | 6/78<br>(7.7%) | 3.3, 16.1 | $p=1.0$ | 6/38<br>(15.8%) | 7.1, 30.8 | $p=0.151$ | 1/16<br>(6.3%) | 0.0, 30.3 | $p=1.0$ |
| MARV GP |  |  |  |  |  |  |  |  |  |  |  |  |  |  |
| Visit 0 | 42/295<br>(14.2%) | 10.7, 18.7 | 25/184<br>(13.6%) | 9.3, 19.3 | $p=0.9488$ | 16/110<br>(14.5%) | 9.1, 22.4 | $p=1.0$ | 7/56<br>(12.5%) | 5.9, 23.9 | $p=0.8937$ | 3/27<br>(11.1%) | 3.0, 28.9 | $p=0.8741$ |
| Visit 1 | 23/240<br>(9.6%) | 6.4, 14.0 | 18/158<br>(11.4%) | 7.2, 17.4 | $p=0.6801$ | 11/95<br>(11.6%) | 6.4, 19.7 | $p=0.7305$ | 4/45<br>(8.9%) | 3.0, 21.3 | $p=0.3507$ | 5/23<br>(12.5%) | 9.2, 42.3 | $p=0.1466$ |
| Visit 2 | 13/172<br>(7.6%) | 4.4, 12.6 | 9/123<br>(7.3%) | 3.7, 13.5 | $p=1.0$ | 10/78<br>(12.8%) | 6.9, 22.2 | $p=0.2724$ | 4/38<br>(10.5%) | 3.6, 24.7 | $p=0.7806$ | 2/16<br>(12.5%) | 2.2, 37.3 | $p=0.8294$ |
| MARV VP40 |  |  |  |  |  |  |  |  |  |  |  |  |  |  |
| Visit 0 | 13/295<br>(4.4%) | 2.5, 7.5 | 13/184<br>(7.1%) | 4.1, 11.8 | $p=0.2975$ | 6/110<br>(5.5%) | 2.3, 11.6 | $p=0.8576$ | 3/56<br>(5.4%) | 1.3, 15.2 | $p=1.0$ | 2/27<br>(7.4%) | 1.0, 24.5 | $p=0.8172$ |
| Visit 1 | 11/240<br>(4.6%) | 2.5, 8.1 | 10/158<br>(6.3%) | 3.3, 11.4 | $p=0.594$ | 5/95<br>(5.3%) | 2.0, 12.0 | $p=1.0$ | 4/45<br>(8.9%) | 3.0, 21.3 | $p=0.4104$ | 3/23<br>(13.0%) | 3.7, 33.0 | $p=0.2149$ |
| Visit 2 | 4/172<br>(2.3%) | 0.7, 6.0 | 3/123<br>(2.4%) | 0.5, 7.2 | $p=1.0$ | 4/78<br>(5.1%) | 1.6, 12.8 | $p=0.4361$ | 4/38<br>(10.5%) | 3.6, 24.7 | $p=0.0546$ | 1/16<br>(6.3%) | 0.0, 30.3 | $p=0.9037$ |

